# Identification of novel causative genes of childhood epileptic encephalopathies

**DOI:** 10.1101/2023.07.25.23293037

**Authors:** Yi-Wu Shi, Jian-Guo Zhang, Zi-Long Ye, Na He, Wen-Bin Li, Han-Kui Liu, Zhi-Gang Liu, Nan-Xiang Shen, Xiao-Chong Qu, Cui-Xia Fan, Jie Wang, Sheng Luo, Yu-Dan Lv, Li Gao, Jing Chen, Shao-Ping Huang, Xin-Guo Lu, Jing Yu, Jie Zhang, Xiu-Xia Wang, Qu-Wen Gao, Li Jiang, Yan-Hui Chen, Hui Qi, Jing-Da Qiao, Li-Zhi Chen, Yuan-Jin Zeng, Xiao-Xue Yang, Hong-Jun Yan, Cheng-Yan Li, Tao Zeng, Fu-Li Min, Bing Qin, Hai-Qing Xu, Lin Xu, Bing-Mei Li, Yong-Hong Yi, Zhi-Hong Zhuo, Qing-Hui Guo, Su-Li He, Hong-Wei Zhang, Li-Ping Guan, Wei-Yi Deng, Xiao-Fan Ren, Dong-Fang Zou, Wei-Yue Gu, Tao Su, Xiao-Rong Liu, Yi-Bo Qu, Xin-Ping Yang, Wei-Ping Liao

**Author notes:** Correspondence to Dr. Liao, Department of Neurology, Institute of Neuroscience, Key Laboratory of Neurogenetics and Channelopathies of Guangdong Province and the Ministry of Education of China, The Second Affiliated Hospital, Guangzhou Medical University, Guangzhou, China,;. These authors contributed equally to the manuscript (Yi-Wu Shi, Jian-Guo Zhang, Zi-Long Ye).

## Abstract

**Background and Objectives:** Epileptic encephalopathy is a devastating epilepsy with etiologies largely elusive, despite recent whole gene/exon sequencing on large cohorts. This study targeted on the genetic causes of childhood epileptic encephalopathy, typically Lennox-Gastaut syndrome (LGS) featured by age-dependent onset and characteristic clinical manifestations.

**Methods:** Trio-based whole-exome sequencing was performed in 241 unrelated cases from 21 epilepsy centers through China Epilepsy Gene 1.0 Project with individualized analyses on each trio by explainable inheritance origin with stratified frequency filtration, on each gene in four aspects that include gene expression in the brain, previously reported phenotypes, pLI/pRec/Z-missense index, and genetic knockout/knockdown phenotypes, followed by multiple/specific statistical analyses depending on the inheritance pattern of variants, including the establishment of controls for analysis of compound heterozygous variants.

**Results:** Three genes presented highly repetitive variants with statistical significances, including *SBF1* with *de novo*, *CELSR2* with recessive, and *TENM1* with X-linked recessive variants. The frequency of compound heterozygous/homozygous *CELSR2* variants in the cases was significantly higher than that in 1942 asymptomatic parent controls. The variants located at functional domains in *SBF1* and *TENM1*, and homozygous/compound heterozygous variants with one of the paired variants located in functional domains in *CELSR2*, were associated with more frequent and refractory seizures. Six genes were suggested to be potential causative genes with evidence of gene profile and statistical significance. Additional 41 genes were considered as potential candidate genes with evidence of gene profile/statistical significance.

**Discussion:** This study suggests that *SBF1*, *CELSR2*, and *TENM1* are causative genes of LGS and highlights the implication of individualized analysis protocol, i.e., individualization on phenotype, individualization on each case, and individualization on each gene, in identifying genetic causes of human diseases.

## Introduction

Developmental epileptic encephalopathies (DEEs) represent a clinically and genetically heterogeneous group of devastating epilepsies characterized by refractory seizures, severe electroencephalography (EEG) abnormalities, and neurodevelopmental delay or decline^1^. It is a common clinical entity and accounts for approximately 40% of newly diagnosed epilepsies before the age of three years^2^. DEE differs in the age of onset and contains early infantile epileptic encephalopathy with onset in neonates or infants^3^, such as infantile epileptic spasms syndrome and Dravet syndrome, and childhood-onset epileptic encephalopathy, typically Lennox-Gastaut syndrome (LGS)^4^. LGS is estimated to account for 1%-2% of all patients with epilepsy and up to 10% of patients with childhood epilepsy^4–6^. It usually begins between 1 and 8 years old (before age 18 years) and is characterized by multiple seizure types that must include tonic seizures and/or consistent paroxysmal fast activity/polyspike on EEG^4^. The etiologies of epileptic encephalopathies remain largely elusive. Recent studies have demonstrated an increased number of genes associated with epileptic encephalopathies. To date, 112 genes have been confirmed to be linked to DEEs (https://omim.org), explaining 22% to 40% of cases with epileptic encephalopathy of neonates or infants^7,8^. In contrast, just a limited number of genes like *CHD2*, *DNM1*, *GABRB3*, and *SYNGAP1*, have been identified in patients with LGS, accounting for a small proportion of cases^9–12^. Furthermore, recent studies with large cohorts have encountered challenges in discovering novel DEE genes^13–15^. However, the age-dependent feature, specific clinical characteristics, and distinct EEG pattern highly suggest an underlying genetic mechanism in the epileptogenesis of LGS, which still to be determined.

In the present study, we conducted trio-based whole-exome sequencing (WES) on a cohort of patients with LGS who had no acquired etiologies. The potential pathogenic variants were screened by an individualized analysis protocol that includes individualized analyses on each trio by explainable inheritance origin with stratified frequency filtration, on each gene from four aspects that include gene expression in the brain, previously reported phenotypes, probability of being intolerant to heterozygous/homozygous variants of loss-of-function or missenses (pLI/pRec/Z-missense), and genetic knockout/knockdown phenotypes, followed by statistical analyses depending on the inheritance pattern of variants. We utilized a control group consisting of 1942 asymptomatic parents to directly analyze the frequency of compound heterozygous variants statistically. *SBF1*, *CELSR2,* and *TENM1* were identified as disease-causative genes and 47 genes were suggested to be potential causative/candidate genes of LGS. This study suggests the implications of the individualized analysis protocol in identifying genetic causes of human diseases, which will be useful for future genetic medicine.

## Materials and Methods

### Subjects

We recruited 241 unrelated LGS patients (240 trios and 1 adopted singleton) without acquired causes from 21 hospitals (Table S1) through the China Epilepsy Gene 1.0 Project (C-epg1.0; https://epg1.cn), which comprises 16 epilepsy sub-cohorts stratified by clinical phenotype. All subjects were of Han Chinese ethnicity, with all four grandparents being Han Chinese and parents having no blood relatives.

Comprehensive clinical data were collected, including basic information, seizure details, treatment response, family history, and examination results. Brain MRI examinations were conducted to detect structural irregularities in the brain. Patients with acquired etiologies were excluded. Video EEG assessments were conducted, and their outcomes were evaluated by two certified electroencephalograph specialists. The diagnosis and classification of epileptic seizures and LGS followed the criteria of the International League Against Epilepsy (1989-2025). All enrollees were diagnosed with LGS, with frequent seizures characterized by two or more of the following features: (1) multiple seizure types with tonic seizures among them, (2) generalized polyspikes or fast rhythms during sleep (especially required when daily tonic seizures are obscure), and (3) generalized slow (< 2.5 Hz) spike-wave complex on EEG. Each patient was under follow-up for a minimum of twelve months.

This research complied with the patient consent guidelines for research participation outlined by the International Committee of Medical Journal Editors and obtained approval from the ethics review board of the Second Affiliated Hospital of Guangzhou Medical University (approval ethics number 2020-hs-49). The patients themselves or their legal representatives provided written informed consent.

### Whole-exome sequencing

Genomic DNA was isolated from the peripheral blood samples of patients and their parents (trios) via the QIARamp Blood Mini Kit. Trio-based WES was carried out on a HiSeq 2000 platform as previously reported^12,16^. Sequencing data from massive parallel sequencing had average depth > 100× and > 98% coverage of captured regions. High-quality reads were aligned to GRCh37 via Burrows-Wheeler alignment, with single-nucleotide variants and insertions/deletions identified using Genome Analysis Tool Kit.

### Variant evaluation

We developed an individualized analytical framework that includes five steps (Fig. 1).

1. General filtration. Variants with potential pathogenicity, such as initiation codon, nonsense, frameshift, canonical splice site, in-frame insertion/deletion, missense, and synonymous variants predicted to affect splicing, were kept. Subsequently, these variants underwent filtering to retain only those with a minor allele frequency (MAF) under 0.005 in the Genome Aggregation Database (gnomAD).
2. Variant filtration by explainable inheritance origin in each trio. The genetic origin of variants reflects the genetic differences between the affected individual and their parents, thereby accounting for the phenotypic manifestations in a specific family (trio), i.e., *de novo,* recessive (each inherited from the one asymptomatic parents), or hemizygous. Variants with an explainable inheritance origin in each trio were selected.
3. The variants were then filtered with stratified MAF criteria. For *de novo* variants, hemizygotes, and homozygotes, the MAF is set to be absent in control populations in gnomAD. For compound heterozygous variants, the product of the two alleles frequencies in gnomAD is < 1×10^-6^, a value sevenfold lower than the probability of an individual in the current gnomAD population (1/141456 = 7 × 10^-6^). The criteria are relatively strict because variants that confer risk for phenotypes that decrease fecundity (such as LGS) are necessarily rare in the population since they are transmitted less frequently than neutral alleles, which is a result of negative selection. In contrast, the filtering MAF for mild phenotypes should be individualized to be relatively high, such as the *GJB2*-associated deafness, in which the two hotspot recessive variants (c.109G>A/p.Val37Ile and c.101T>C/p.Met34Thr) present MAFs of 0.007556 and 0.008996, respectively^17^.
4. Filtration based on the gene profile, using data for each gene summarized in Genetic Dependence & Pathogenicity Database (GDP; www.gdap.org.cn). Genes with qualified variants were classified into epilepsy-associated genes (977 genes by Wang *et al.*^18^ and updated from OMIM) and genes with undefined gene-disease/epilepsy/seizures associations. The pathogenicity of variants in epilepsy-related gene was evaluated in accordance with the guidelines and standards of the American College of Medical Genetics and Genomics (ACMG)^19^. For genes of undefined gene-disease/epilepsy/seizure associations, the following four aspects were assessed: (1) Tissue-specific expression: a candidate causative gene of epilepsy ought to be expressed in the brain (inclusion criterion), with careful consideration of the possibility of other accountable pathogenic mechanisms, such as ectopic expression and distant toxic effects of abnormal metabolic products. (2) Excluding the disease-causing genes with pre-established gene-disease associations (genotype-phenotype correlation) where epilepsy is not considered a possible phenotype (exclusion criterion). (3) Consideration was given to the probability of intolerance to heterozygous/homozygous loss-of-function/missense variants (pLI/pRec/Z-missense), including genes with *de novo* variants and pLI ≥ 0.9, genes with recessive variants and pLI ≥ 0.9/pRec ≥ 0.9/pNull ≤ 0.1, and genes with missense variants and Z-missense ≥ 3.1. (4) Whether the genetic knockout/knockdown produces phenotypes of the brain. These four aspects determine or are closely related to gene-disease (epilepsy) association.
5. Multiple/specific statistical analyses. The Poisson cumulative distribution function^9^ was calculated in genes with *de novo* variants. For genes with biallelic variants, we used the cumulative binomial probability^20^ and established a control cohort to directly analyze the frequency of compound heterozygous variants statistically. Additionally, the aggregate frequency of variants^21^ was calculated for each gene. For excess *de novo* variants, mutation rate of each gene was determined by summing point mutation rates in effectively captured consensus coding regions among trio offspring and dividing by total trios (240 in this cohort). The *P*-value was computed as [1-Poisson cumulative distribution function (χ-1, λ)], with χ denoting the observed number of *de novo* variants for a specific gene and λ calculated as 2×240×1.2×10^-8^×effective transcript length of the gene on the autosome^9^. For excess biallelic variants, the *P*-value was computed as [1-cumulative binomial probability [(n-1, N, R)], with n representing observed count of biallelic variants for a specific gene, N stands for number of trios (240 in this cohort), and R denotes the random variant frequency of the gene within the population^20^. The variants were filtered by different MAF cutoffs based on the class of variants, i.e., biallelic loss-of-function (LOF), one LOF and one damaging missense, and biallelic damaging missense. The threshold was set based on the MAF in the ExAC-East Asian population, given that all patients were Han Chinese. For the aggregated frequency of variants, the control populations include the East-Asian controls and all gnomAD controls. It is currently unknown the frequencies of compound heterozygous variants in general populations. To assess the significance of the compound heterozygous variants, we constructed a control cohort consisting of 1942 asymptomatic parents from trios, in which the compound heterozygous variants in parents were identified by detecting one of the paired variants in their offspring since one of the paired variants from each parent would be transmitted to the child.

**Fig. 1.**
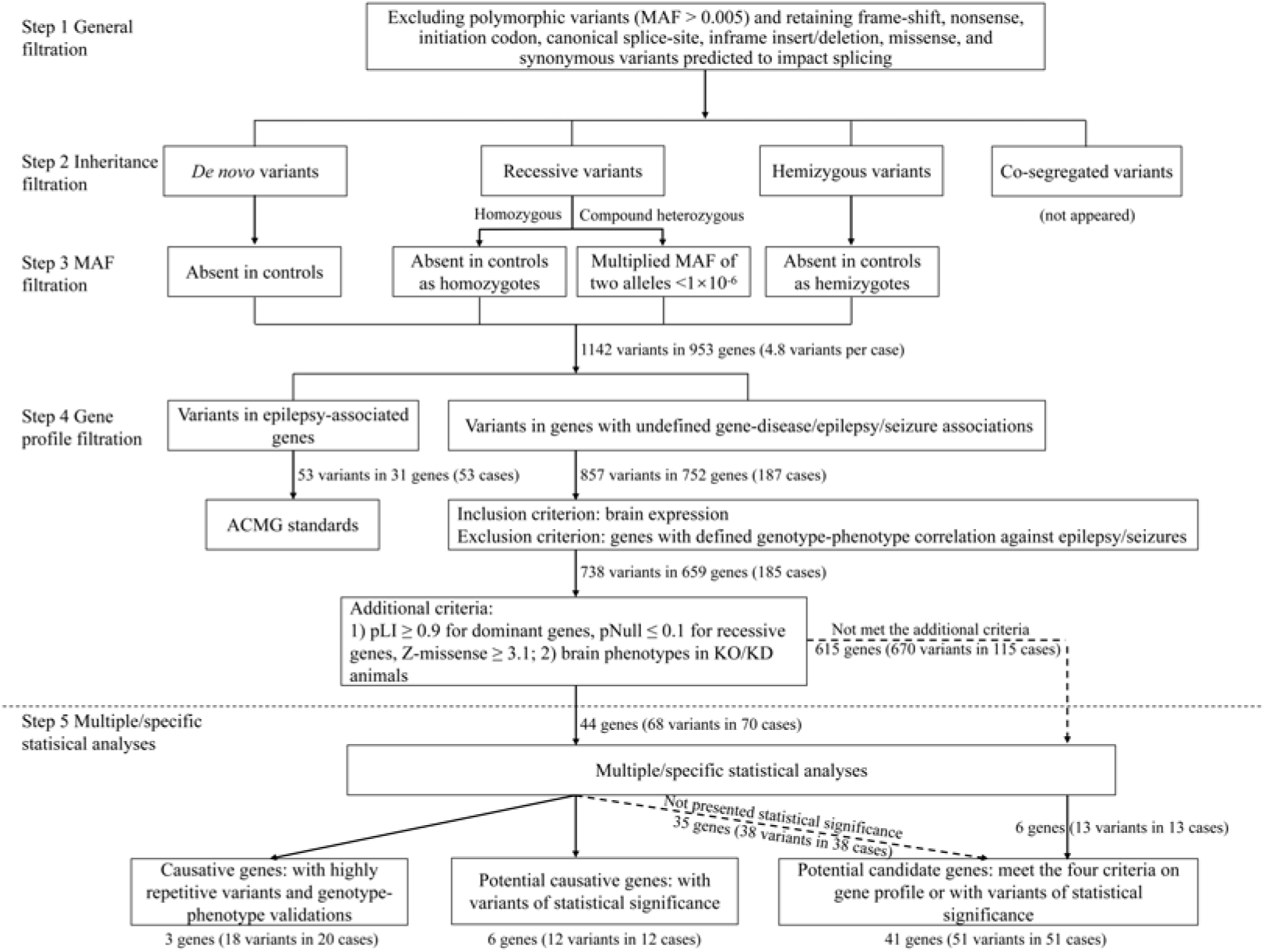
Flow chart of trio-based whole-exome sequencing with an individualized analytical framework. In the step 3, the MAFs in the control populations in gnomAD were used for filtrations. For compound heterozygous variants, the product of multiplying the frequencies of two alleles in gnomAD is < 1×10^-6^, which is 7 times less than the probability of one individual in the population of gnomAD (1/141456 = 7 × 10^-6^). Abbreviations: ACMG, American College of Medical Genetics and Genomics guidelines; KO/KD, knockout/knockdown; MAF, minor allele frequency.

Finally, the potential disease-associated genes were classified into disease-causative, potential causative, and potential candidate genes, based on evidence of different levels (Fig. 1). Genes met all criteria and with genotype-phenotype validations are considered as disease-causative genes. Genes met the four criteria on gene profile and with variants of statistical significance are considered as potential causative genes. Genes met the four criteria on gene profile or with variants of statistical significance are considered as potential candidate genes.

All candidate variants were confirmed via Sanger sequencing. Qualified novel candidate genes were subjected to further bioinformatics and experimental studies to establish the associations between genes and diseases.

### Protein structure modeling

The tertiary structure of the proteins produced by the novel candidate genes was modeled using trRosetta (https://yanglab.nankai.edu.cn/trRosetta/) to predict how missense variants affect protein structure. Hydrogen bonding was analyzed and visualized via PyMOL 2.3 software.

### Statistical analyses

The allele frequencies between the case and control groups were compared using the two-tailed Fisher’s exact test. *P* values less than 0.05 were considered statistically significant. Analysis of the excess of *de novo* and biallelic variants in a specific gene was performed according to previous reports^9,20^ (detailed above in **Variant evaluation**). Bonferroni correction for *P* values was applied for analysis of the excesses of variants (*Pc* < 0.05/19,711, given the general count of encoding genes in the human genome). All testing was conducted analyzed by using GraphPad Prism 8.2.

## RESULTS

The present LGS cohort recruited 240 unrelated trios and a singleton (adopted) from 21 hospitals (Table S1), which was from the C-epg1.0 Project that consisted of 16 sub-cohorts of epilepsy, including 4 sub-cohorts of DEE.

The 241 recruited patients included 169 boys and 72 girls, and the average age of onset was 4.6 years old (4.6 ± 3.8, mean ± SD). On average, 5.9 Gb of the sequence was produced within the exome-targeted regions in each individual, with an average coverage of 120 depths (Table S2).

To identify candidate causative variants, we adopted an individualized analytical framework that includes individualized analysis of each trio by explainable inheritance origin with stratified frequency filtration and individualized analysis of each gene from four aspects, followed by statistical analyses depending on the inheritance pattern of the variants (Fig. 1).

After variant filtration by inheritance origin and stratified minor allele frequency (MAF) from gnomAD database (steps 2 and 3 in Fig. 1), we obtained 1142 qualified variants (4.8 per case), including 180 single *de novo* (0.8 per case), 78 homozygous (0.3 per case), 746 compound heterozygous (3.1 per case), and 138 hemizygous variants (0.8 per male) (Fig. 2 and Table S3). Individualized analysis of each gene was then performed with separation of epilepsy-associated genes and genes of unknown pathogenic potential.

**Fig. 2.**
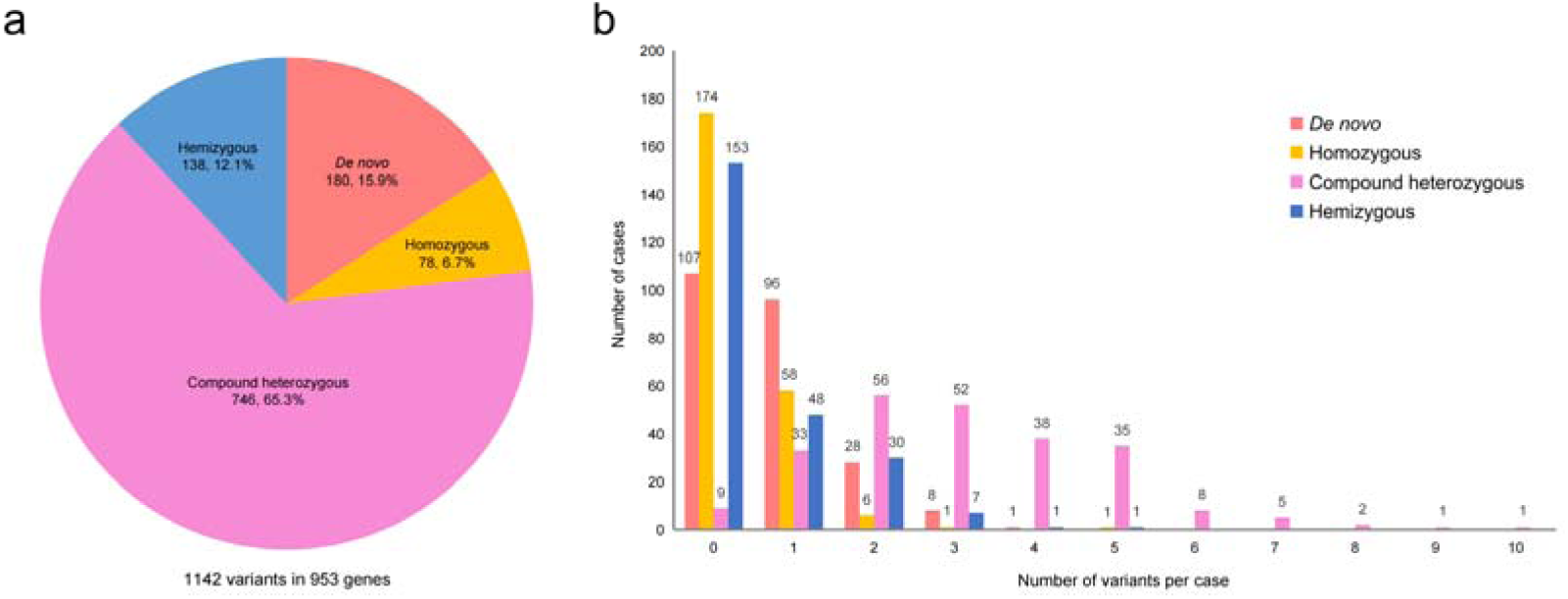
General information of the variants after filtration by inheritance origin and stratified MAF. **(a)** Pie chart of the inheritance origin of the 1142 qualified variants. **(b)** The distribution of the number of *de novo*, homozygous, compound heterozygous, and hemizygous variants per case.

### Potential pathogenic variants in epilepsy-associated genes

Variants in epilepsy-associated genes were identified in 53 cases with 31 genes involved (Fig. 1, Table 1 and S4). Among these variants, 47 single *de novo* variants in 26 genes were evaluated as pathogenic or likely pathogenic by using ACMG^19^ standards, explaining 19.5% of the cases (47/241) in this cohort. Variants in *CHD2* and *SETD1B* showed significant excesses of *de novo* variants^9^ (Table S5), explaining 5.8% of the cases (14/241).

**Table 1.**
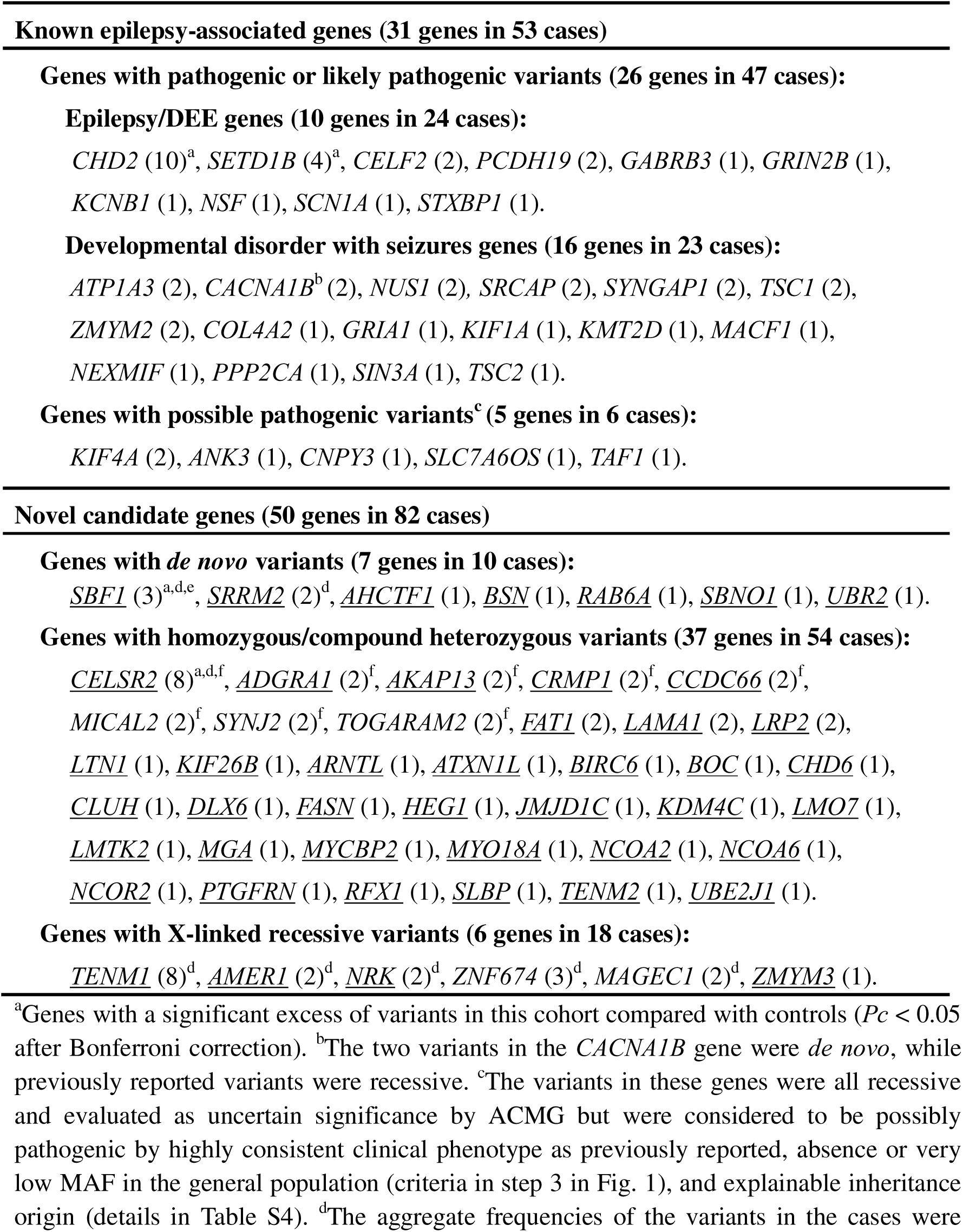
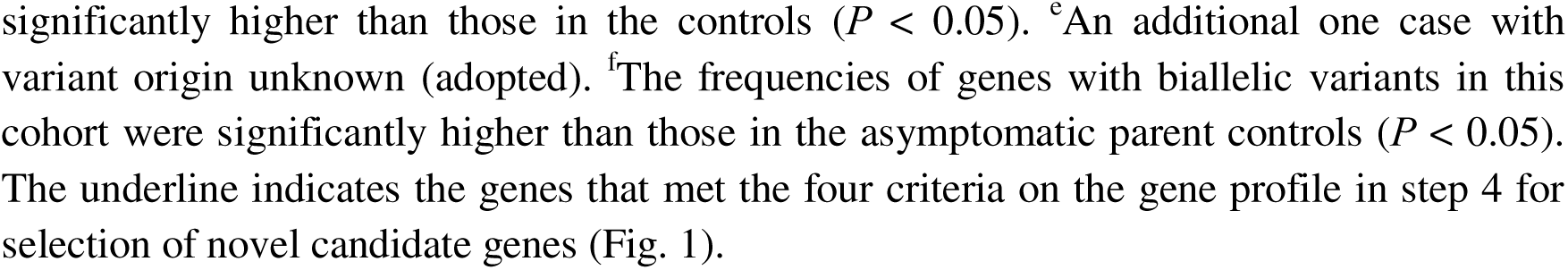
Overview of potential causative genes in this cohort Known epilepsy-associated genes (31 genes in 53 cases)

It was noted that the pathogenic/likely pathogenic variants estimated by ACMG are all of *de novo*. In contrast, 6 recessive variants were evaluated as uncertain significance by ACMG (Table 1 and S4), including 2 homozygous, 1 compound heterozygous, and 3 hemizygous variants. Given that ACMG evaluates each of the paired biallelic variants separately and does not consider the specific dose effect of hemizygous variants, the pathogenicity of these variants was re-evaluated. These variants were considered to be possibly pathogenic by highly consistent clinical phenotype as previously reported, absence or very low MAF in the general population (criteria in step 3 in Fig. 1), and explainable inheritance origin (Table S4).

### Potential pathogenic variants in novel candidate genes

For the variants in genes of unknown pathogenic potential in each family (trio, 187 in total), analyses on gene profile were performed, which referred 857 variants (752 genes). This process was facilitated by using the dataset summarized in the GDP Database. After initial analyses on gene profile, 93 genes that did not meet the criterion of inclusion/exclusion were excluded, and 609 genes that passed the criterion of inclusion/exclusion but did not meet the two additional criteria were evaluated as uncertain significance (Fig. 3). Forty-four genes presented evidence from four aspects of the gene profile, which substantially supported the gene-disease association (Fig. 1, at the end of Step 4).

**Fig. 3.**
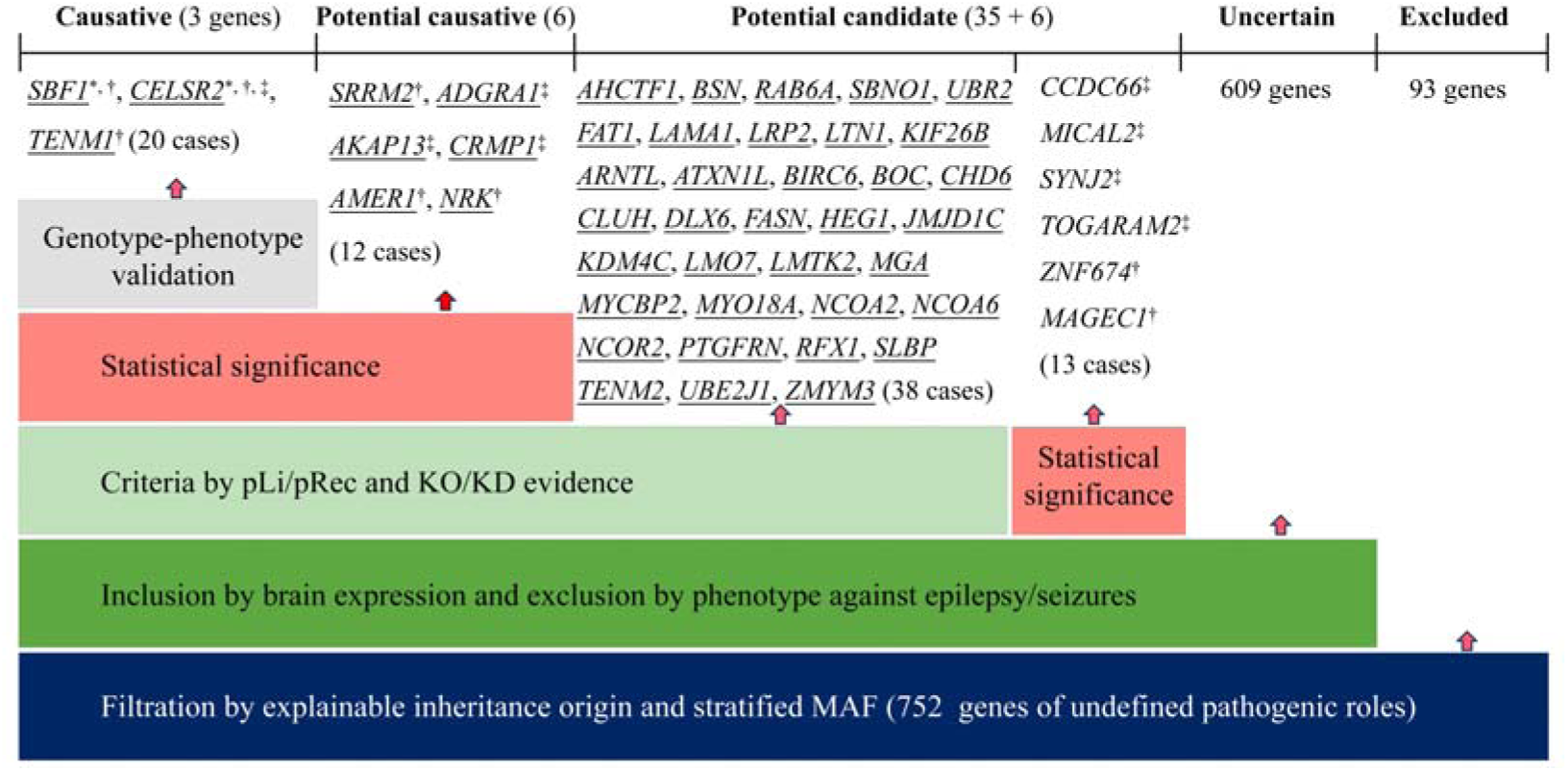
Targeting and defining novel causative genes by individualized analyses. This includes individual analyses on each trio by explainable inheritance origin with stratified frequency filtration, on each gene from four aspects that include tissue-specific expression, previously reported phenotypes, probability of being intolerant to heterozygous/homozygous variants of loss-of-function or missenses (pLI/pRec/Z-missense), and KO/KD phenotypes, followed by multiple/specific statistical analyses depending on the inheritance pattern of variants. The potential disease-associated genes were classified into disease-causative, potential causative, and potential candidate genes, based on evidence of different levels. Abbreviations: KO/KD, knockout/knockdown; MAF, minor allele frequency. The underline indicates the genes that met the four criteria on the gene profile. *, genes with a significant excess of variants in this cohort compared with controls (*Pc* < 0.05 after Bonferroni correction); †, the aggregate frequencies of the variants in the cases were significantly higher than those in the controls (*P* < 0.05, Table S9); ‡, the frequencies of genes with biallelic variants in this cohort were significantly higher than those in the asymptomatic parent controls (*P* < 0.05, Fig. 4).

After statistical analyses (Fig. 3, and Table 1), nine genes met the criteria of gene profile and showed statistical significance, suggested to be potentially causative with 32 cases involved (Fig. 3, Table S6 and S7). *SBF1*, *CELSR2*, and *TENM1* appeared highly repetitively (in four or more cases) and were validated to be disease-causative by genotype-phenotype correlation.

*SBF1* missense variants appeared in four cases (three *de novo* and one unknown origin variants, Table S6), which presented significant excess of *de novo* variants by calculating the Poisson cumulative distribution function^9^ (significant after Bonferroni corrections, *Pc* = 0.0095, Table S5).

*CELSR2* biallelic variants appeared in eight cases (Table S6), which presented significant cumulative binomial probability for recessive variants^20^ (1% MAF cutoff for biallelic damaging missense genotypes, 8 versus 0.0029, *Pc* = 0.014). To analyze the significance of biallelic variants, we established a control cohort of compound heterozygous variants derived from 1942 asymptomatic parents. The frequency of compound heterozygous variants in the cases was significantly higher than that in the asymptomatic parent controls (8/241 vs 0/1942, *P* = 1.99 × 10^-8^; Fig. 4 and Table S8). In contrast, *SPTBN5* compound heterozygous variants appeared in five cases, but also appeared 15 times in the controls, being not statistically significant (5/241 vs 15/1942, *P* = 0.061; Fig. 4 and Table S8).

**Fig. 4.**
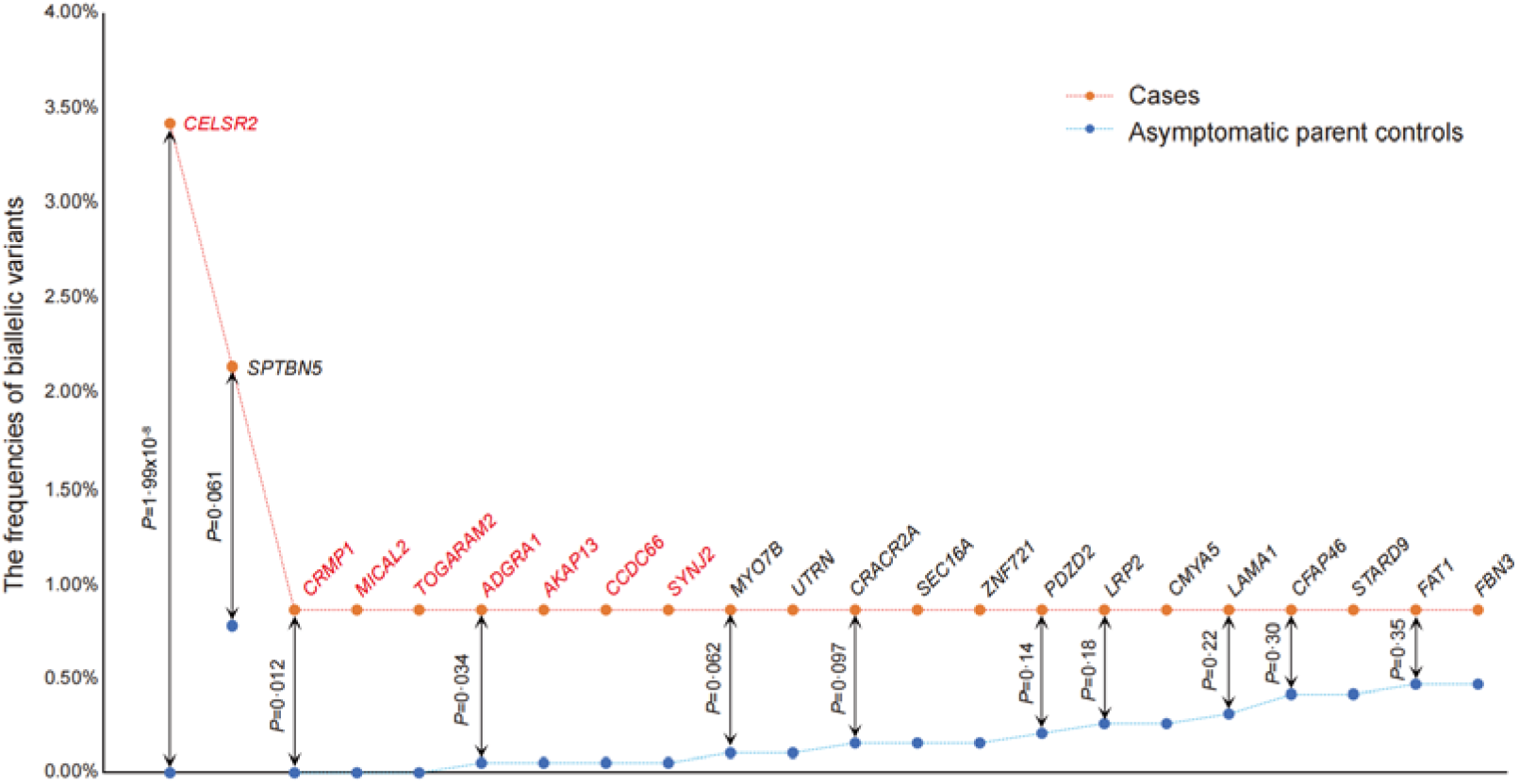
The frequencies of biallelic variants in LGS compared with those in the asymptomatic parent controls. The orange dots indicate the frequencies of genes with biallelic variants in cases. The blue dots indicate the frequencies of genes with biallelic variants in asymptomatic parent controls (detailed numbers in Table S8). The genes in red font indicate that the frequencies of biallelic variants in cases were significantly higher than those in the asymptomatic parent controls. Data were analyzed by two-tailed Fisher’s exact test.

*TENM1* hemizygous variants were identified in 8 unrelated cases (7 variants including a repeated variant, Table S6), which was significantly more frequent than the male controls (Table S9). The variants in *SBF1* and *CELSR2* also presented higher aggregate frequencies of the variants in the cases than those in the specific controls (Table S9).

Additional six genes passed the criterion of inclusion/exclusion and presented statistical significance, be considered as potential candidate genes with 13 cases involved (Fig. 3 and Table S10).

### Genetic-clinical features of the patients with variants in *SBF1*, *CELSR2*, and *TENM1*

Detailed information was provided in Table 2.

**Table 2.**
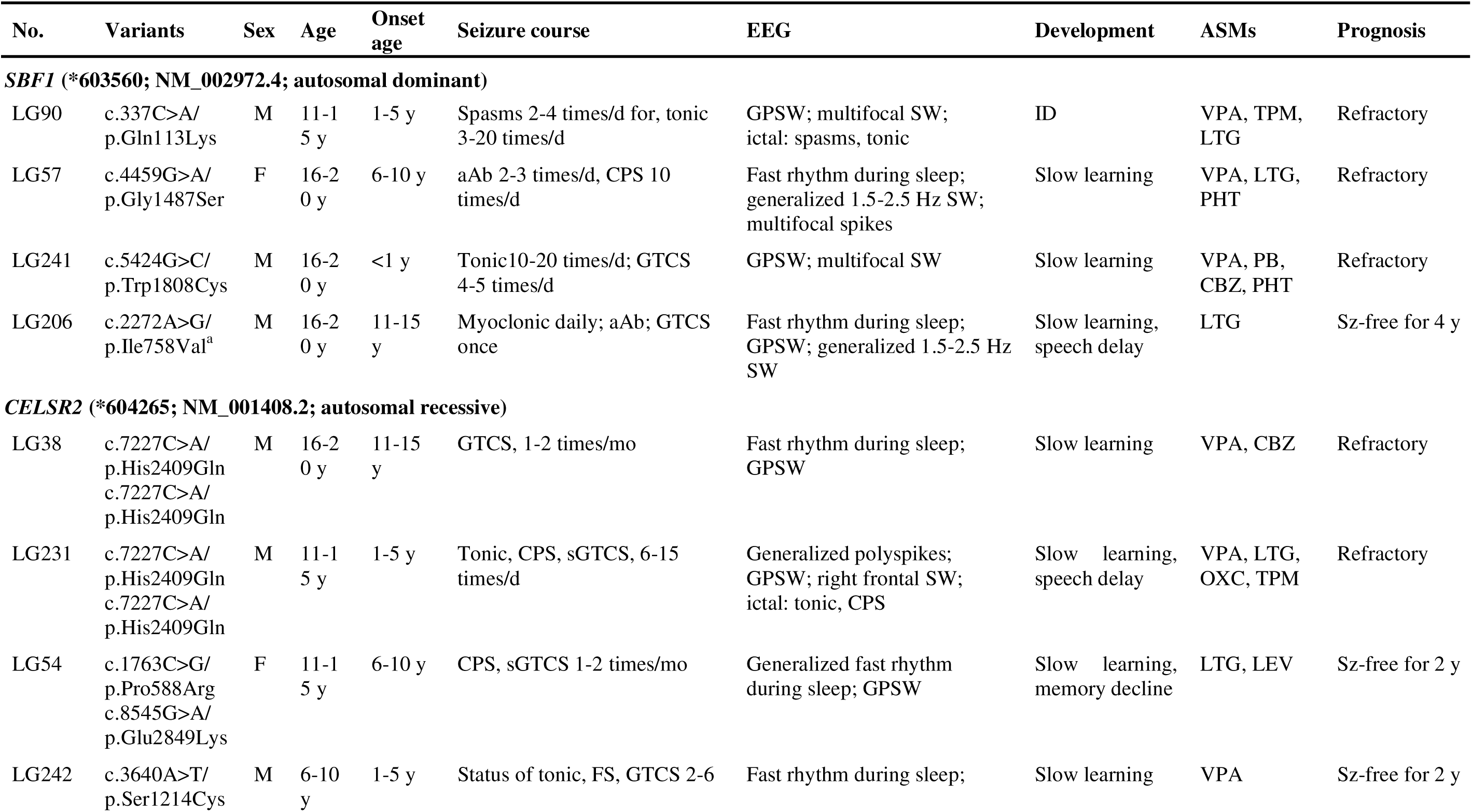
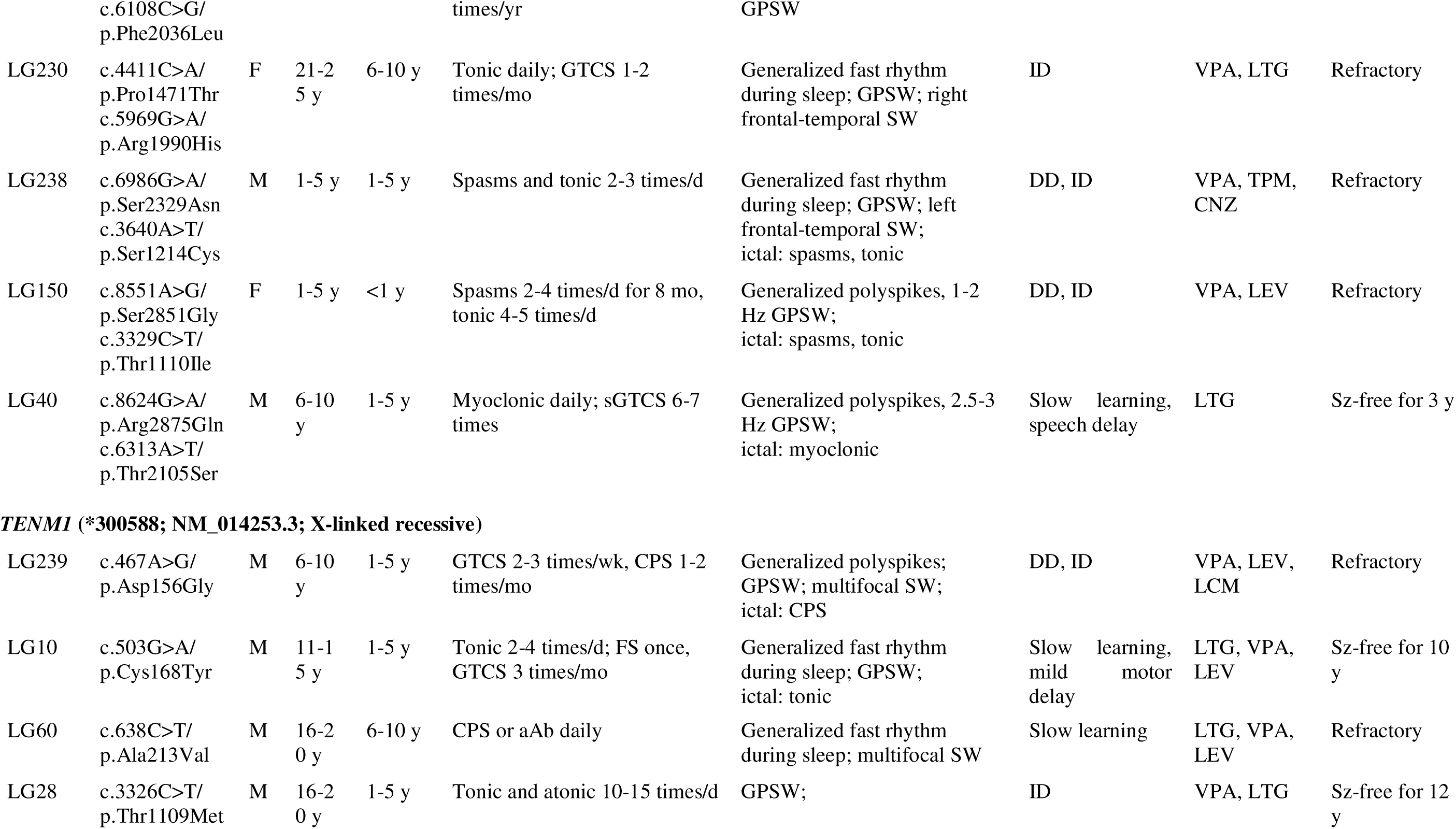
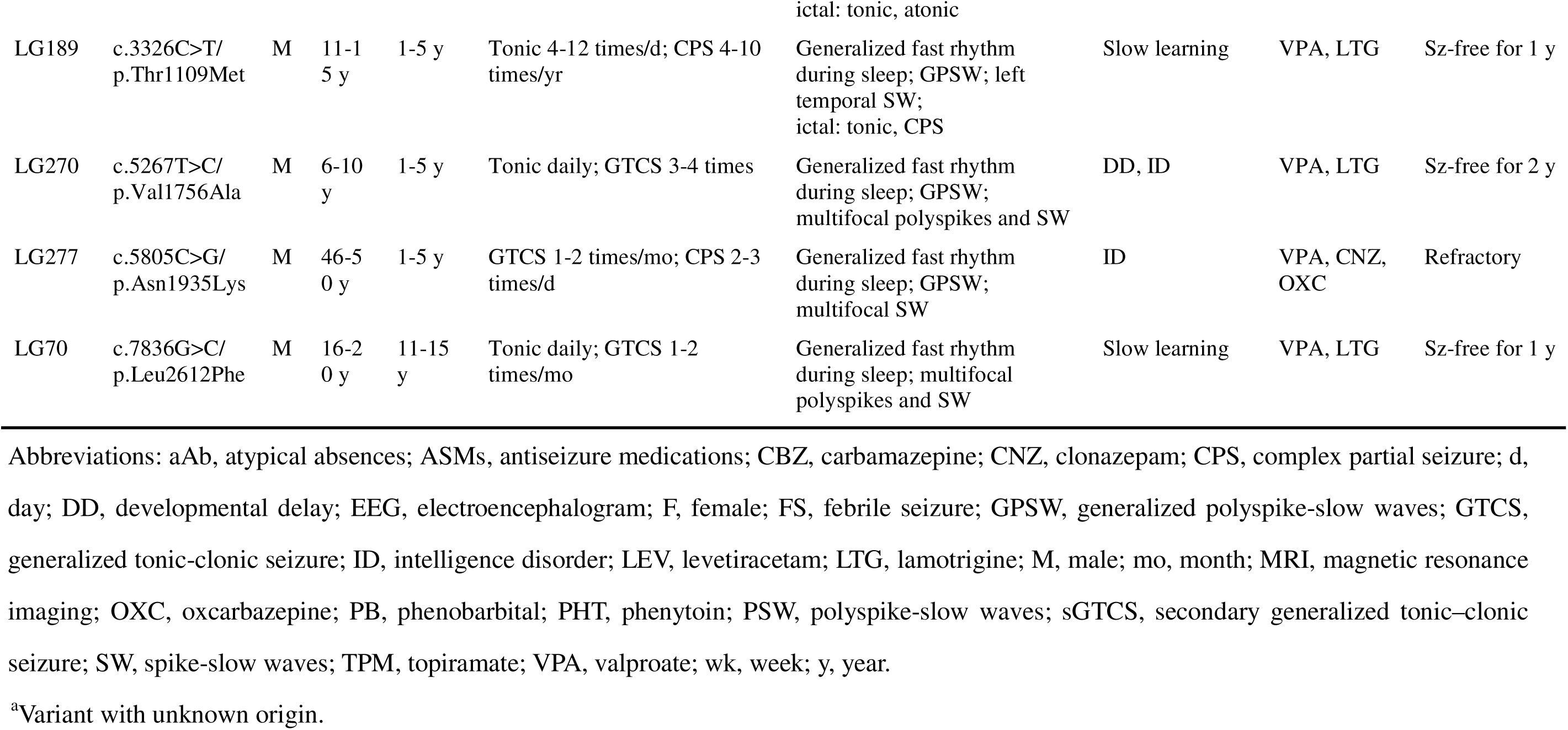
Clinical features of the patients with variants in *SBF1*, *CELSR2*, and *TENM1*.

### SBF1

Four missense variants in *SBF1* were identified in four unrelated cases, including three *de novo* variants (c.337C>A/p.Gln113Lys, c.4459G>A/p.Gly1487Ser, and c.5424G>C/p.Trp1808Cys) and one with unknown origin (adopted, c.2272A>G/p.Ile758Val) (Fig. 4a and the DNA sequencing chromatograms in Fig. S1a). The four variants were absent in the controls of gnomAD-all population, and exhibited a significant excess of *de novo* variants (Table S5) and significantly higher aggregated frequencies than those in the controls of gnomAD populations (Table S9).

Three *de novo* missense variants (p.Gln113Lys, p.Gly1487Ser, and p.Trp1808Cys) were situated within the functional domains (the tripartite differentially expressed in normal and neoplastic cells (DENN) domain, the myotubularin phosphatase domain, and the pleckstrin homology domain, respectively, Fig. 5a). These variants were predicted to bring about changes in hydrogen bonding with nearby residues (Fig. S1b). The patients (LG90, LG57, and LG241) with intractable and frequent seizures even under a combination of three or four antiseizure medications (ASMs). They presented both generalized and multifocal discharges in EEGs.

**Fig. 5.**
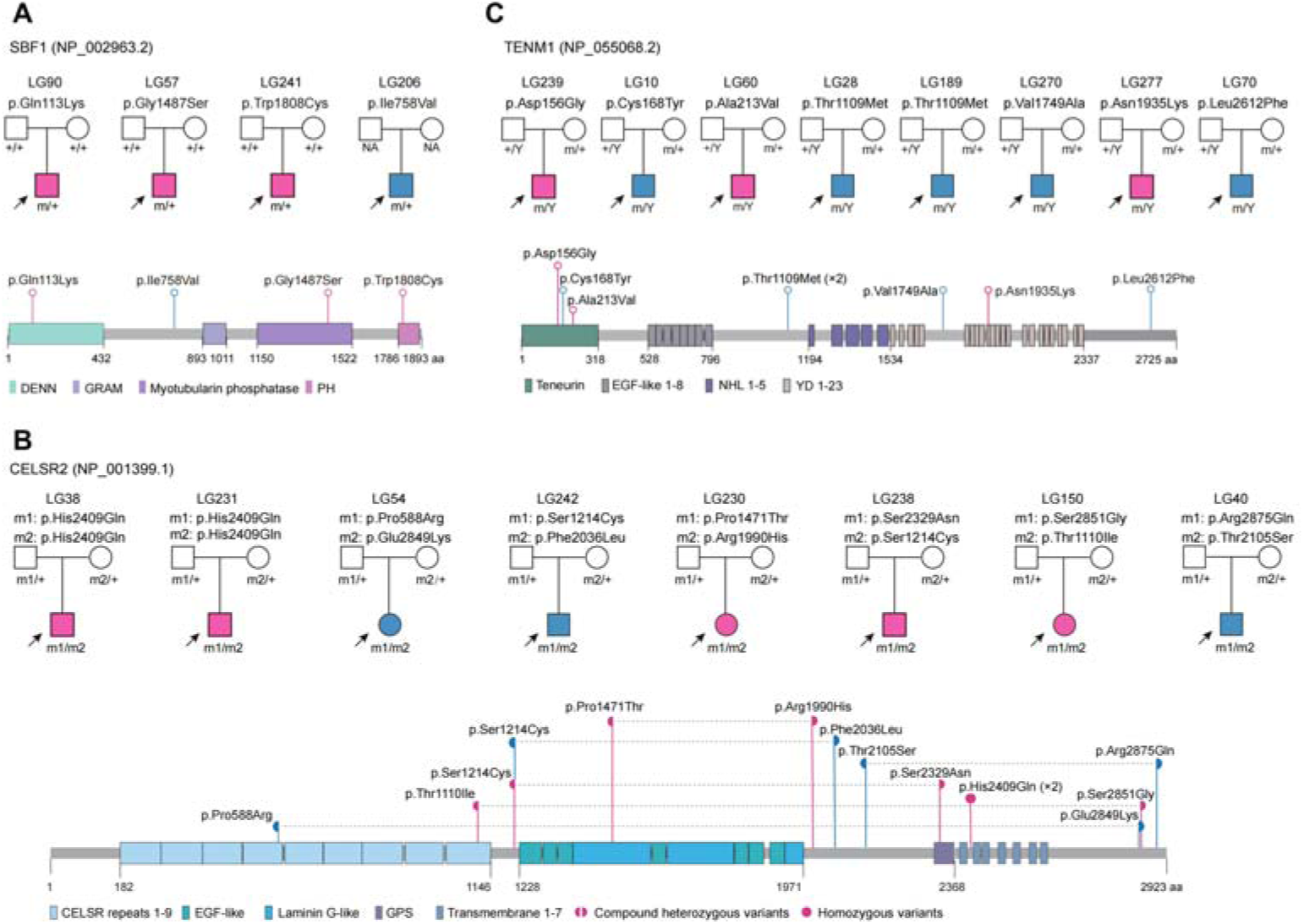
*SBF1*, *CELSR2*, and *TENM1* variants identified in patients with LGS. Pedigrees of the unrelated cases with *SBF1* (a), *CELSR2* (b), and *TENM1* (c) variants (top) and the localization of the variants in the protein domains (bottom). Red-colored individuals (or variants) were associated with refractory epilepsy; and blue-colored individuals achieved seizure-free. All patients experienced frequent seizures of multiple types. The gray dotted lines in (b) indicate a pair of compound heterozygous variants of *CELSR2*. Abbreviations: CELSR, cadherin EGF LAG seven-pass G-type receptor; DENN, differentially expressed in normal and neoplastic cells; EGF, epidermal growth factor; GPS, G-protein-coupled receptor proteolytic site; GRAM, glucosyltransferases, rab-like GTPase activators and myotubularins; NHL, ncl-1, HT2A and lin-41; PH, pleckstrin homology; YD, Tyr and Asp dipeptide.

The variant p.Ile758Val was located in the region between the DENN domain and the glucosyltransferases, rab-like GTPase activators and myotubularins domain without hydrogen bond changes by protein modeling, yet it was predicted to reduce the stability of protein (Fig. S1b). The patient (LG206) presented a relatively late age of onset (11-15 years) with only generalized discharges in EEG. He achieved seizure-free with monotherapy of lamotrigine, despite previous daily myoclonic seizures.

### CELSR2

Biallelic variants in *CELSR2* were identified in eight unrelated cases, including a homozygous missense variant (c.7227C>A/p.His2409Gln) in two cases (Fig. 5b and the DNA sequencing chromatograms in Fig. S2a). The variant p.Ser1214Cys was also identified repeatedly in two cases. These variants exhibited significant excess of biallelic variants, significantly higher aggregated frequencies of variants (Table S9), and significantly higher frequency of compound heterozygous variants (Fig. 4).

Four of the twelve missense variants were located at the functional domains, including two (p.Pro588Arg and p.Thr1110Ile) in the cadherin EGF LAG seven-pass G-type receptor repeat domains, one (p.Pro1471Thr) in the laminin G like domain, and one (p.Ser2329Asn) in the G-protein-coupled receptor proteolytic site domain. The other eight variants were located outside the known-functional domains (Fig. 5b). Among the eight pairs of biallelic variants, at least one of the paired missense variants was predicted to affect hydrogen bonding or decrease the protein stability (Fig. S2b).

The two cases with homozygous variant p.His2409Gln (LG38 and LG231) exhibited intractable seizures. In contrast, the two cases (LG242 and LG40) with compound heterozygous variants that had both variants located outside the known-functional domains (p.Ser1214Cys/p.Phe2036Leu and p.Thr2105Ser/p.Arg2875Gln) achieved seizure-free with monotherapy of valproate/lamotrigine. Among the other four cases with one of the paired variants located in the functional domains, three cases (LG230, LG150, and LG238, who had variants p.Pro1471Thr, p.Thr1110Ile, and p.Ser2329Asn in functional domains, respectively) presented refractory seizures; and the another case (LG54) achieved seizure-free under bi-medication of lamotrigine and levetiracetam, who had compound heterozygous variants with two variants located furthest apart (p.Pro588Arg and p.Glu2849Lys, a distance of 2261 amino acids), which have been suggested to be associated with phenotype severity^22^.

### TENM1

Seven hemizygous missense variants in *TENM1* were identified in eight unrelated cases (Fig. 5c and the DNA sequencing chromatograms in Fig. S3a). All *TENM1* hemizygous variants were inherited from their asymptomatic mothers, in line with an XLR inheritance pattern. No hemizygotes/homozygotes of the variants were present in the controls of gnomAD-all population, and the aggregate frequencies of the variants in this cohort were significantly higher than those in the male controls of gnomAD (Table S9).

Among the three cases with variants in the N-terminal intracellular teneurin domain, two cases (LG239 and LG60) suffered from refractory seizures even under three ASMs; one case (LG10) exhibited frequent seizures but he achieved seizure-free with three ASMs of valproate, lamotrigine, and levetiracetam, who had variant (p.Cys168Tyr) of less hydrophobicity changes (0.58) than that of LG239 (0.77) and LG60 (0.91) (Fig. S3c). The patient (LG277) with variant in Tyr and Asp dipeptide 10 domain (p.Asn1935Lys) exhibited refractory seizures despite a combination of three ASMs (Table 2). In contrast, the four cases (LG28, LG189, LG270, and LG70) with variants located outside the known-functional domains achieved seizure-free under bi-medication of valproate and lamotrigine.

## DISCUSSION

Despite the wide application of high-throughput sequencing, the gene-disease associations in more than three-fourths of the genes in the human genome remain undetermined (https://omim.org). Previous studies on DEE focused on *de novo* variants and identified several disease-causative genes^11,23,24^. However, recent studies on rare/ultra-rare variants are limited in discovering novel DEE-associated genes^13–15^. The present study focused on LGS and collected 241 unrelated cases from 21 epilepsy centers. Although modest in size, it was the largest cohort of LGS so far, to our knowledge. By using trio-based whole-exome sequencing with the individualized analysis protocol, three disease-causative genes, six potential causative genes, and 41 potential candidate genes were identified. LGS is a classical devastating epileptic encephalopathy. The identification of causative genes implies clinical significance in diagnosis and potentially further in management of LGS. This study highlights the implications of individualized analysis protocol, i.e., individualization on phenotype, individualization on each case, and individualization on each gene (3I), in identifying genetic causes of human diseases, which is potentially helpful in identifying novel causative genes among the three-fourths of the human genome.

Epilepsy syndromes are characterized by age-dependent clinical manifestations, especially the age of onset^1^, which are potentially associated with distinct pathogenic genetic causes^3,4,18^ and determined by their genetic dependent stages^25,26^. The present study is on the cohort of LGS, an epileptic encephalopathy of childhood onset with characteristic clinical manifestations, differing from most of the DEEs that are commonly of early onset. Given that more than 100 genes are associated with DEE (https://omim.org), each of the DEE-associated genes may appear in single or few cases of a cohort. Among the 31 defined epilepsy-associated genes, only *CHD2* and *SETD1B* appeared in more than two cases in this study. In fact, some classical causative genes of epilepsy, such as *KCNT1*^13,14^ and *TSC2*^13–15^, appeared rarely in large cohort and may thus be missed by common statistical analyses. The phenotype subclassification, or individualization on phenotype in another word, is therefore critical for identification of specific disease-causative genes, especially for complex clinical entities like epilepsy, as suggested in several of our recent studies^16,26,27^.

This study employed an individualized analysis protocol to screen potential pathogenic variants, including individualized analyses on each trio by explainable inheritance origin with stratified frequency filtration and on each gene from four aspects.

Filtrations by inheritance pattern and MAF have been generally used in clinical genetic tests for genes with defined gene–disease associations, but not commonly used in studies on large cohorts to identify novel disease-associated genes, except that on *de novo* variants^9,28^. In this study, variants from genes of undetermined pathogenic potential were sorted by inheritance in each trio, which represents the genetic difference between the affected and normal individuals (explains the origin). The variants were then filtered with MAF according to the inheritance pattern, instead of a criterion like “absence” in the control populations, which is especially critical for identification of recessive disease-associated genes. Since the human genome is diploid and usually a portion of functioning from one copy of a gene is sufficient for the biophysiological function, pathogenic recessive variants, especially compound heterozygous variants, are potentially common^29–31^. However, since each of the paired compound heterozygous variants is not pathogenic thus potentially presents with low MAF in general populations, typically p.Arg208Ter (MAF of 0.00025) and c.509-1G>C (MAF of 0.00039) in *TPP1* that were identified in 89% of patients with ceroid lipofuscinosis^32^. This brings difficulties to variant filtration and statistical analysis; and criteria of rare/ultra-rare without consideration in inheritance pattern potentially leads to missing the pathogenic recessive variants. Besides cumulative binomial probability^20^, this study established a control cohort for direct statistical analysis of the frequency of compound heterozygous variants, which was suggested to be useful in application by identifying *CELSR2,* but not *SPTBN5* that appeared in five patients, as a disease-causative gene in this study.

The individualized analyses on each trio by explainable inheritance origin with stratified frequency filtration helped to screen variants and avoided missing variants of potentially pathogenic. However, an average of 4.8 variants per case (1142 variants in 953 genes in total) were obtained. Further analyses on gene profile were performed to select and target specific disease-associated genes. Forty-four genes presented evidence from four aspects of the gene profile, which substantially supported the gene-disease association. These evidences include high expression in the brain, high pLI/pRec/Z-missense scores, brain abnormalities in knockout models, and exclusion of the possibility of previously defined gene-disease associations against epilepsy as a phenotype. The individualized analysis protocol in this study analyzed the sequencing data for each trio once the data was available. It is time-consuming and becomes applicable with the help of the dataset summarized in the GDP database (www.gdap.org.cn) and the APP tools.

This study identified 50 candidate genes included six genes with additional statistical evidence and three genes with highly repetitive variants.

*SBF1* encodes a member of the protein-tyrosine phosphatase family, which plays a vital role in cell growth and differentiation^33^. Previously, *SBF1* biallelic recessive variants, including three biallelic null variants (Fig. S4), were reported to be associated with Charcot-Marie-Tooth (CMT) type 4B3, which is characterized by progressive limb muscle weakness and distal sensory impairment^34^. In contrast, *SBF1* variants identified in this study are *de novo* heterozygous missense variants. *SBF1* is expressed in the brain and muscle at a similar level. The pLI of *SBF1* is 1.00, indicating that *SBF1* is highly intolerant to heterozygous loss-of-function variants. Additionally, the difference between the missense Z-score and the synonymous Z-score is 7.54, indicating that *SBF1* is high intolerance of missense variants. These evidences suggest a potential association between epilepsy and heterozygous *SBF1* missense variants. Recent studies have suggested that the abnormal proteins associated missense variants play a critical role in the pathogenic mechanism of epilepsy^26,35,36^. The mechanism underlying the pathogenesis of *SBF1* biallelic variants in CMT4B3 is unclear^37^. Genes associated with both CMT and epilepsy have been previously reported, such as *AARS1*^38,39^ and *DYNC1H1*^40–42^, for which further studies are needed to determine the underlying mechanisms.

*CELSR2* encodes a transmembrane protein of the flamingo subfamily, which is highly expressed in the brain and plays a vital role in cell/cell signaling during nervous system formation^43^. Currently, *CELSR2* have not been defined to be associated with human diseases (https://omim.org). Our previous studies have shown that two CELSR subfamily genes, *CELSR1* and *CELSR3*, are associated with epilepsy^44,45^. *TENM1* encodes a transmembrane teneurin subfamily protein, which is highly expressed in the brain and participates in neural development by regulating the formation of appropriate connectivity within the nervous system^46,47^. At present, there is no report on *TENM1*-associated disease. The pLIs of both *CELSR2* and *TENM1* are 1.0, with high missense Z-score (3.91 for *CELSR2* and 3.43 for *TENM1*), indicating that both genes are highly intolerant to heterozygous loss-of-function and missense variants.

Clinically, potential genotype-phenotype correlations were observed in each of *SBF1, CELSR2,* and *TENM1*. The variants located at functional domains in *SBF1* and *TENM1*, homozygous variants or compound heterozygous variants with one of the paired variants located in functional domains in *CELSR2*, and variants with more hydrophobicity change of amino acid residue in *TENM1*, were associated with more frequent and refractory seizures. All the 20 patients with variants of these genes presented frequent seizures of multiple types; but nine of them achieved seizure-free with timely treatment (particularly valproate and/or lamotrigine), which were all explained by the genotype-phenotype correlations. LGS is a typically childhood-onset epileptic encephalopathy characterized by frequent and refractory seizures. The identification of causative genes and the disclosed genotype-phenotype correlations potentially imply a significance in the diagnosis and management of the patients with LGS.

## Conclusions

This study identified three novel causative genes of childhood epileptic encephalopathy, *SBF1* with *de novo* dominance inheritance, *CELSR2* with biallelic recessive inheritance, and *TENM1* with X-linked recessive inheritance, which disclosed genotype-phenotype correlations are significance in clinical diagnosis and management. This study highlights the implication of individualized analysis protocol in combination with specified statistical analysis and setting controls for compound heterozygous variants in identifying genetic causes of human diseases.

## Data Availability

All data produced in the present study are available upon reasonable request to the authors

## Data availability

The data that support the findings of this study are available from the corresponding author upon reasonable request.

## Acknowledgements

We thank the affected patients and their families for participating in this study. This work was funded by the National Natural Science Foundation of China (grant nos. 82171439, 82271505, and 81971216) and the Guangdong Basic and Applied Basic Research Foundation (grant no. 2021A1515010986 and 2022A1515111123). The funders had no role in the study design, data collection, and analysis or in the decision to publish or the preparation of the manuscript.

## Author Contributions

WPL and YWS conceptualized the study, analyzed and interpreted the data, and drafted and revised the manuscript. WPL had full access to all data in the study and takes responsibility for the integrity of the data and accuracy of the data analysis. JGZ, ZLY, NH, NXS, and HKL performed the clinical data and whole-exome sequencing analysis. WPL, YWS, JGZ, ZLY, NH, NXS, HKL, WBL, XCQ, and CXF drafted and revised the manuscript and contributed to the statistical analysis. All authors collected data, revised the manuscript, and contributed to the writing.

## Ethics statement

All procedures performed were in accordance with the ethical standards of the institutional committee. Ethical approval was approved by the ethics committee of the Second Affiliated Hospital of Guangzhou Medical University (approval ethics number 2020-hs-49). We declared that all patients’ IDs were not identified to anyone outside the research group.

