## Supplemental Figures for "Identification of novel causative genes of childhood epileptic encephalopathies"

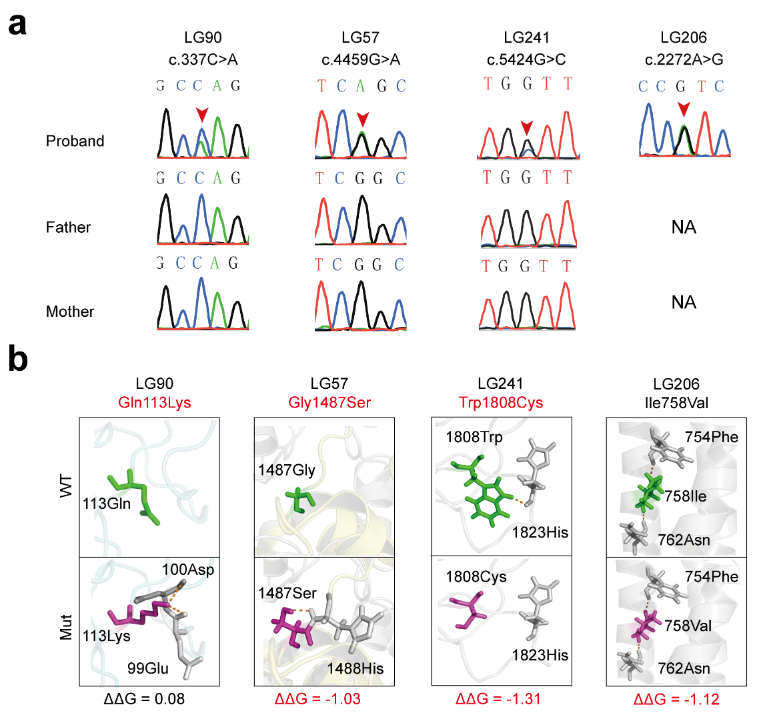

**Fig. S1. Genetic data of cases with *SBF1* variants. (a)** Sanger sequencing verification of four cases with *SBF1* variants. **(b)** Modeling of *SBF1* missense variants indicated abnormal hydrogen bonds. Wild-type and mutant residues are colored green and light magenta, respectively. Hydrogen bonds are colored orange. The variant Lys113 rebuilds two hydrogen bonds with Asp100 and Glu99. Ser1487 rebuilds one with His1488. The variant Cys1808 destroyed one hydrogen bond with His1823. Variants that altered the stability of the global conformation of the SBF1 protein were indicated by the value of free energy stability changes (ΔΔG, Kcal/mol) using online tools (<http://gpcr.biocomp.unibo.it/cgi/predictors/I-Mutant3.0/I-Mutant3.0.cgi>). An absolute value of ΔΔG > 0.5 indicates a large decrease in stability. The variants in red font (the top) indicate that the variant changed hydrogen bonds with surrounding amino acid residues. The ΔΔG in red font (bottom) indicates that the variant significantly changed the protein stability.

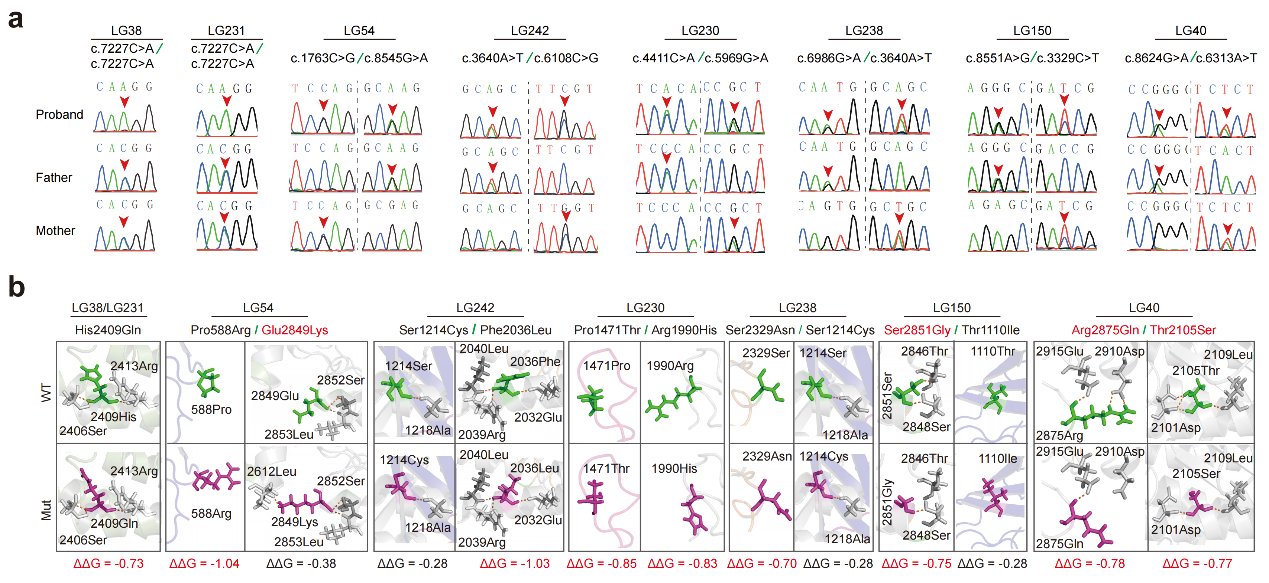
 **Fig. S2. Genetic data of cases with *CELSR2* variants. (a)** Pedigrees and Sanger sequencing verification of the eight cases with biallelic *CELSR2* variants. **(b)** Modelling of CELSR2 missense variants indicated abnormal hydrogen bonds. Wild-type and mutant residues are colored green and light magenta, respectively. Hydrogen bonds are colored orange. Variants that destabilized the global conformation of the CELSR2 protein were indicated by the value of free energy stability changes (ΔΔG, Kcal/mol) using online tools (http://gpcr.biocomp.unibo.it/cgi/predictors/I-Mutant3.0/I-Mutant3.0.cgi). An absolute value of ΔΔG > 0.5 indicates a large decrease in stability. The variants in red font (the top) indicate that the variant changed hydrogen bonds with surrounding amino acid residues. The ΔΔG in red font (bottom) indicates that the variant significantly changed the protein stability.

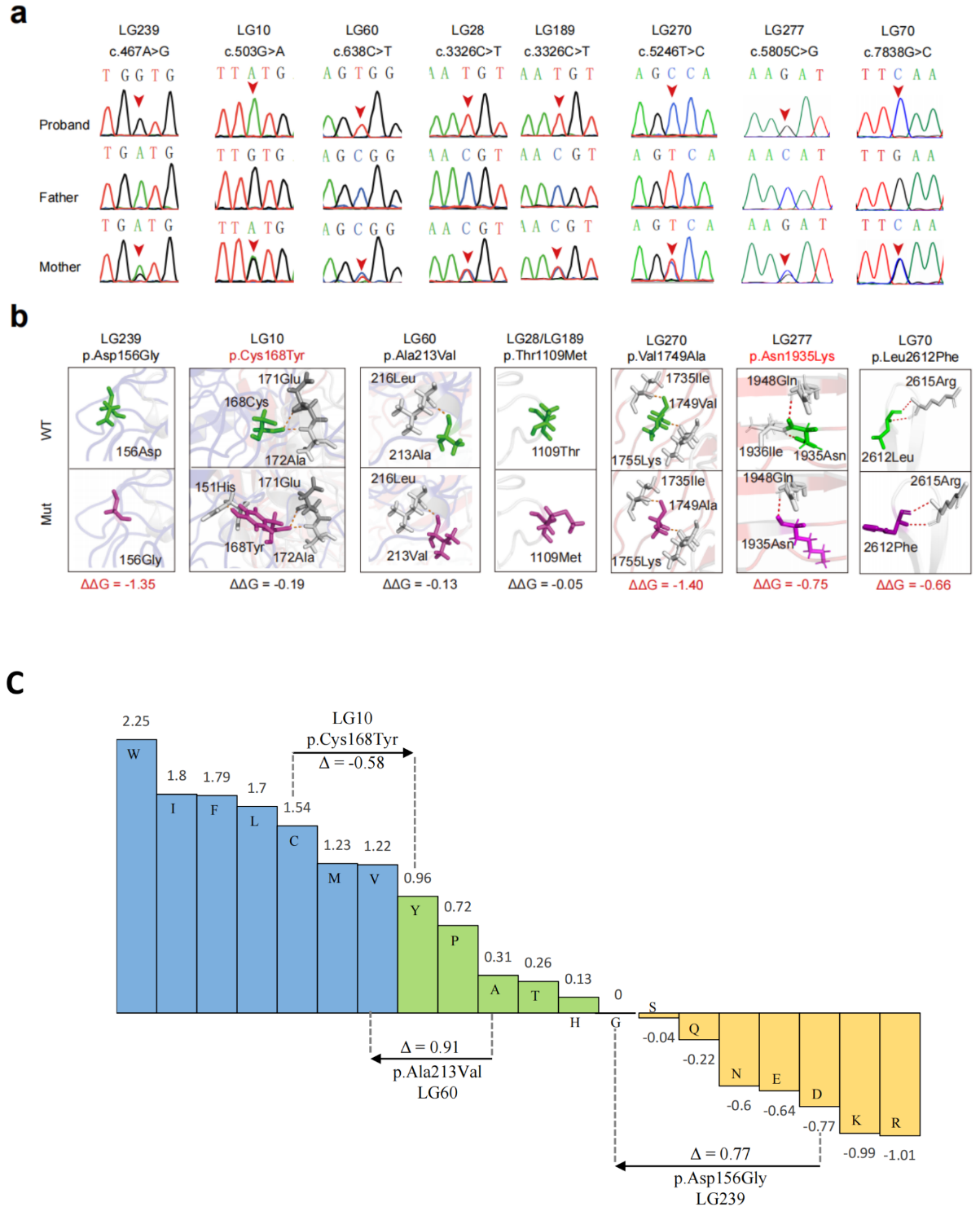
 **Fig. S3. Genetic data of cases with *TENM1* variants. (a)** Pedigrees and Sanger sequencing verification of the six cases with *TENM1* variants. **(b)** Modelling of TENM1 missense variants predicted abnormal hydrogen bonds. Wild-type and mutant residues are colored green and light magenta, respectively. Hydrogen bonds are colored orange. The variant Tyr168 rebuilds a hydrogen bond with His151. The variant Asn1935 destroyed one hydrogen bond with Ile1936. Variants that destabilized the global conformation of the TENM1 protein were indicated by the value of free energy stability changes (ΔΔG, Kcal/mol) using online tools (http://gpcr.biocomp.unibo.it/cgi/predictors/I-Mutant3.0/I-Mutant3.0.cgi). An absolute value of ΔΔG > 0.5 indicates a large decrease in stability. The variants in red font (the top) indicate that the variant changed hydrogen bonds with surrounding amino acid residues. The ΔΔG in red font (bottom) indicates that the variant significantly changed the protein stability. **(c)** Fauchère and Pliska hydrophobicity scale exhibited the hydrophobicity of 20 amino acids. Abscissa: from left to right, hydrophobicity gradually decreased. Blue amino acids are hydrophobic, green amino acids are neutral, and yellow amino acids are hydrophilic. Amino acids with high positive values are more hydrophobic, whereas amino acids with low negative values are more hydrophilic. Among the three cases with variants in the N-terminal intracellular teneurin domain, two cases (LG239 and LG60) suffered from refractory seizures even under three ASMs; one case (LG10) exhibited frequent seizures but he achieved seizure-free with three ASMs, who had variant (p.Cys168Tyr) of less hydrophobicity changes (0.58) than that of LG239 (0.77) and LG60 (0.91).

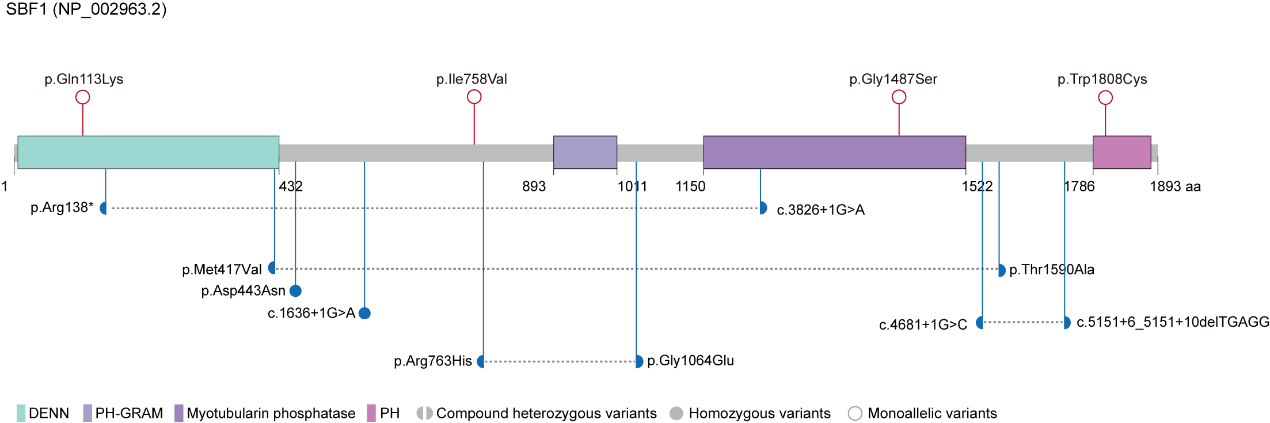
 **Fig. S4. Localization of *SBF1* variants identified in this study and in Charcot-Marie-Tooth** **disease.** Variants identified in this study are shown in red (the top). Variants associated with Charcot-Marie-Tooth disease are shown in blue (bottom). The gray dotted lines indicate the pairs of compound heterozygous variants of *SBF1*.
